# Long COVID research often lacks controls and does not acknowledge this limitation: meta-epidemiological analysis

**DOI:** 10.64898/2026.05.16.26353381

**Authors:** Antonios-Periklis Panagiotopoulos, Alexandros Laskaris, Deny Tsakri, Yiannis Manoussopoulos, Cleo Anastassopoulou, Athanasios Tsakris, John P. A. Ioannidis

## Abstract

**Objectives:** To quantify the frequency of baseline control-group use in published long COVID prevalence studies and assess their key methodological features.

**Methods:** We performed a meta-epidemiological assessment of 440 post-acute COVID-19 prevalence publications from an existing systematic review. To evaluate study design and methodological transparency, we extracted data on the inclusion and classification of comparator groups, the exclusive use of self-reported outcome measures, and whether uncontrolled investigations explicitly recognized the omission of a control group as a limitation. In addition, we surveyed by email the corresponding authors of these articles to determine if any supplementary comparative data existed. The protocol was prospectively registered (DOI: 10.17605/OSF.IO/T2UP9).

**Results:** Among 440 studies, 372 (84.5%) reported no control group. Healthy or uninfected comparators were reported in 55 studies (12.5%) and other comparator types in 14 (3.2%); 1 study included both categories. Solely self-reported outcomes were used in 279 studies (63.4%). Among 372 uncontrolled studies, 244 (65.6%) did not explicitly acknowledge the absence of a baseline comparator as a limitation. Corresponding authors of 140 studies (31.8%) responded to the survey; 126 (90.0%) reported no additional comparative data, while 14 (10.0%) mentioned some available comparative datasets (19 additional datasets). Almost all that information (10/14, 17/19) had been already published in other articles not captured by the index systematic review. Studies with controls had modestly higher citation impact (median 7 versus 4 per year, p=0.002).

Conclusions

Most published long COVID prevalence studies lacked comparator groups and relied exclusively on self-reported outcomes without acknowledging this limitation. Direct author contact identified little additional comparator information. Much of the long COVID prevalence literature may therefore be poorly suited to estimating burden attributable specifically to SARS-CoV-2.

**Key Points:** *Question:* What is the frequency of baseline control group inclusion and the reliance on subjective outcomes in published long COVID prevalence studies?

*Findings:* This meta-epidemiological analysis demonstrates that over 80% of published long COVID prevalence studies lacked a baseline non-COVID control group. Most of these investigations also relied exclusively on subjective patient-reported outcomes without explicitly acknowledging the absence of a comparator as a limitation.

*Meaning:* These findings suggest that the majority of the long COVID prevalence literature is poorly suited to accurately estimate the symptom burden specifically attributable to SARS-CoV-2.

## 1. Introduction

Post-acute sequelae are an intriguing and debated feature of the SARS-CoV-2 post-pandemic legacy. Recent estimates indicate that as many as 36.3% of infected individuals worldwide may experience long-term symptomatic manifestations [1]. However, it is unclear what percentage of these manifestations are causally specific to and triggered by the SARS-CoV-2 infection. Many people in the general population may have chronic symptoms of varying intensity and persistence due to various conditions. Moreover, chronic, debilitating symptom clusters following an acute infection have been described for many other pathogens [2]. An extensive array of viral, bacterial, and parasitic pathogens has historically been claimed to trigger post-acute infection symptoms [2]. Recent evaluations suggest that individuals recovering from non-SARS-CoV-2 acute respiratory infections frequently report very similar prolonged symptom profiles pointing to a substantial and previously overlooked burden of lingering sequelae mimicking long COVID [3].

Both for COVID-19 and other infections, rigorous data on uninfected controls are essential to differentiate true post-infection effects from background symptoms in the general population. Comparison of the effects of COVID-19 versus other infectious exposures is also essential to understand whether COVID-19 may have a different behavior than other pathogens regarding any post-infectious sequelae. Uncontrolled observational designs and single-arm case series are fundamentally limited by the absence of a baseline [4]. Baseline controls are indispensable but do not guarantee unbiased results, as they may still suffer from other biases [5–7]. However, results may be generally more reliable when controls are used and objective outcomes rather than self-reported symptoms are assessed.

It is currently unknown whether studies that do not report proper control groups in their publications simply lacked the ability to collect such data, or if these data are available but remain unreported in the original publications. To address this critical methodological gap, we conducted a comprehensive meta-epidemiological evaluation of the studies identified by a recent systematic review [1] to objectively quantify the frequency of inclusion of baseline control groups and to assess study design efficacy and methodological transparency. To examine possible non-reporting of available comparator data, we contacted the corresponding authors of these studies regarding the availability of supplementary published or unpublished comparative data on uninfected individuals or patients with other confirmed respiratory infections.

## 2. Methods

### 2.1 Study design

We conducted a meta-epidemiological evaluation of post-acute COVID-19 studies [1]. The study methods, prespecified classifications, data extraction procedures, and author survey protocol were registered prospectively before completion of the author survey at 10.17605/OSF.IO/T2UP9. Additional details, including the survey templates and extraction rules, are provided in the protocol and supplementary material (Supplementary Table 1, Supplementary Material 1, Supplementary Material 2). Reporting followed the specifications of the protocol and there is no available reporting guideline for this type of investigation.

We utilized the comprehensive dataset of 442 articles on post-acute COVID-19 prevalence previously identified through a systematic review [1] that had queried Embase, PubMed, and the Web of Science Core Collection for articles published between July 5, 2021 and July 23, 2024. Eligible investigations involved human subjects with verified COVID-19 diagnosis (via clinical assessment, antibody testing, or polymerase chain reaction) and evaluated long COVID outcomes no less than two months post-diagnosis [1]. We excluded formally retracted studies and those that were not in English language.

### 2.2 Data extraction and classification

Full texts were screened and classified using prespecified criteria. The screening and data extraction workload was divided between two reviewers (A-P.P. and A.L.), with each reviewer assessing a distinct subset of studies. A second extraction stage was subsequently performed to collect data regarding use of self-reported outcomes and the acknowledgment of missing controls. During this second stage, each reviewer (A-P.P. and A.L.) independently re-verified the accuracy of the data extracted from their own assigned subset. Uncertain cases were resolved through discussion between reviewers and, when necessary, consultation with a senior reviewer (C.A.).

We established the availability and precise methodological classification of any comparator or control group used by the investigators. Methodological details were recorded in a standardized electronic database (Supplementary Table 1) according to the following predefined classifications:

- Healthy/Uninfected: This category comprised participants drawn from the general population who were either healthy (asymptomatic community-dwelling population) or asymptomatic-confirmed negative for SARS-CoV-2 (by PCR, serology test, rapid test or clinical evaluation). It also encompassed patients admitted to hospital settings for conditions entirely unrelated to COVID-19, e.g. in orthopedic units.
- Other: This heterogeneous classification encompassed various alternative control methodologies. It included cohorts with non-SARS-CoV-2 viral infections, retrospective intra-subject designs comparing pre-and post-COVID-19 symptoms, and patients presenting with similar clinical respiratory features who ultimately tested negative for SARS-CoV-2 (by PCR, rapid test or clinical evaluation).
- No Control: This classification was assigned to any single-arm observational study that failed to report the use of any comparator group.

We also manually extracted the following parameters from all evaluated studies:

- Self-Reported Outcomes: We evaluated whether the study relied entirely on patient self-reported outcomes and symptoms, or whether outcome assessment also included clinician evaluation, laboratory testing, imaging, or other objective measurements.
- Acknowledged Limitations: For all studies classified as “No Control,” we assessed whether the authors explicitly mentioned the lack of a baseline control group as a constraint in the article’s Discussion section or dedicated limitations section. To enhance the robustness of this extraction, we supplemented our manual review with an automated text-mining pipeline. Available PDF documents were processed by Y.M. using a tool [8] which is built to detect limitations sections, alongside custom R scripts to identify manuscripts entirely lacking a limitations section and to flag control-related terminology throughout the entire text (Supplementary material 3). All algorithmic findings underwent subsequent manual re-verification step by A.-P.P. to eliminate false positives (Supplementary material 4).

### 2.3 Author survey protocol

To systematically investigate the potential existence of additional comparative data, a targeted email survey was deployed to corresponding authors of all screened studies. The specific survey inquiries were customized based on the exact control group classification of each study:

- No Control Group: Authors of these studies were asked if their research team collected any supplementary information regarding individuals that could have formed any control group, as defined above (healthy/uninfected, influenza/other documented pathogen, other).
- Control Group: Authors reporting any type of control group were asked about the availability of supplementary comparison data of any additional control groups, as defined above.

We asked authors to tell us about unpublished comparator group information (Supplementary material 2). However, we recognized in the process that some authors might have published on comparator groups elsewhere in other articles, but these potential control groups had not been included in the evaluated studies recorded by Hou et al. Both unpublished and published-elsewhere groups were considered eligible.

We utilized two different email templates (one for controlled studies and one for uncontrolled studies) and because some corresponding authors had multiple entries across both categories, they received up to two emails (Supplementary material 2). Consequently, an author might reply regarding one of their studies while failing to respond to another. Therefore, the final survey numbers represent responses to individual studies rather than unique authors.

The initial survey invitation was sent between February 25 and 27, 2026. A single follow-up reminder was dispatched between March 6 and 9, 2026. The survey database formally closed on March 19, 2026. When an author’s initial response was ambiguous or required further detail, a supplementary clarification email was dispatched following the formal closure date, providing a final window for a definitive reply before the final data closure on March 29, 2026. To maximize data completeness, initial responses received during the clarification window were also included.

When authors provided additional control datasets, these were treated as separate studies.

### 2.4 Data analysis

Data were summarized using descriptive statistics. Frequencies and percentages were calculated for control-group use, control-group type, outcome assessment method, acknowledgment of uncontrolled design among uncontrolled studies, and author survey responses.

As a post-hoc analysis, we also explored the citation impact of studies with different design features. Citation counts were extracted from Scopus on May 2026. For 25 articles published in journals not indexed by Scopus, citations were retrieved from Google Scholar and subsequently halved to approximate equivalent Scopus metrics (Supplementary material 5). The primary metric utilized was citations per year, calculated by dividing total citations by the time elapsed since publication. Statistical comparisons of annual citation rates between controlled versus uncontrolled designs, and self-reported versus other outcome measurements were conducted using the non-parametric Mann-Whitney U test. Four group comparisons of all combinations of control presence and symptom reporting methods were performed utilizing the Kruskal-Wallis test.

## 3. Results

Two of the 442 studies were excluded from analysis (one formal retraction, one non-English). Consequently, 440 individual studies were retained (Figure 1).

**Figure 1.**
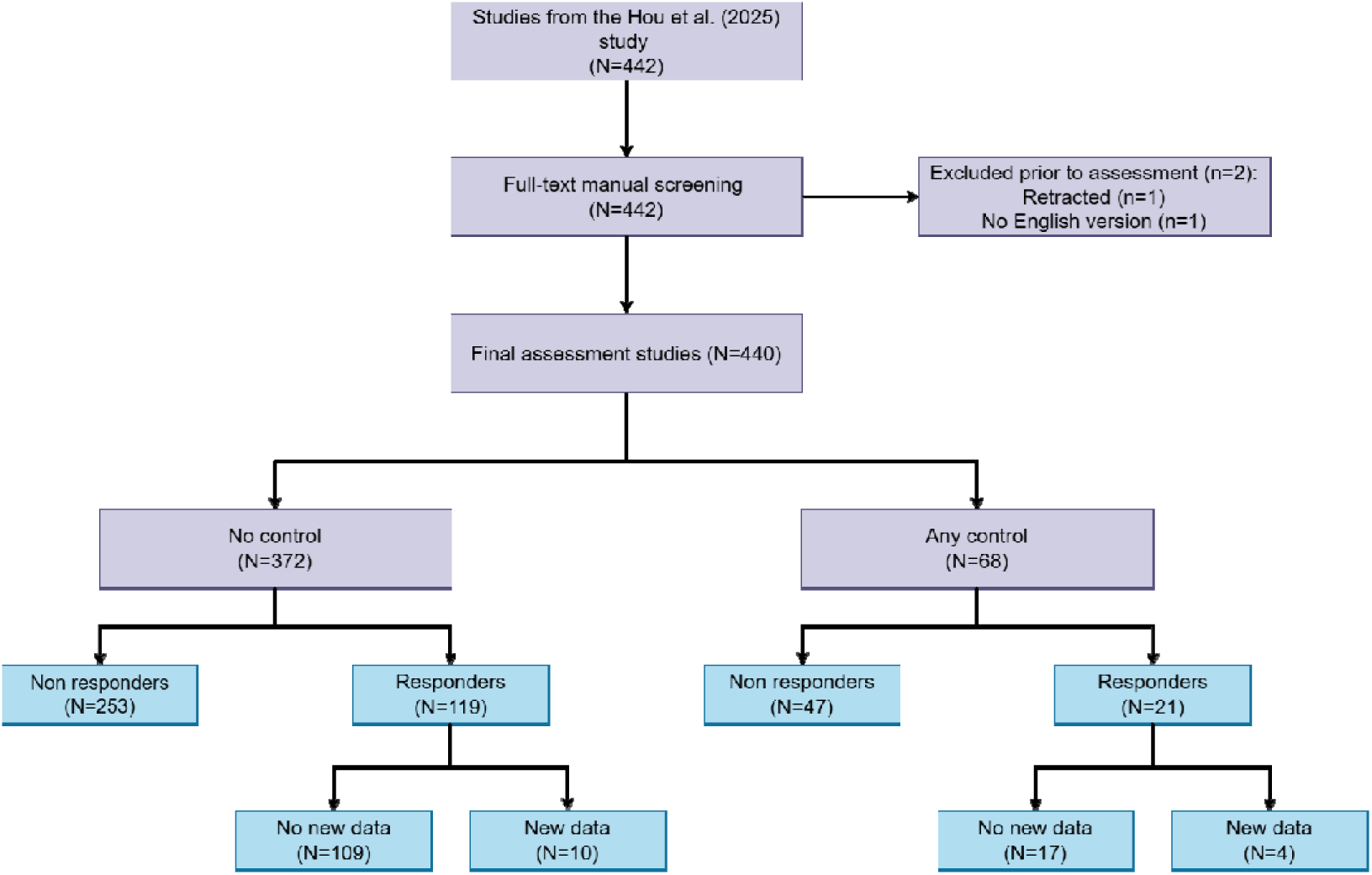
Flowchart of study selection, methodological assessment, and author survey outcomes.

### 3.1 Methodological characteristics of the original cohort

Of the 440 studies analyzed, 372 (84.5%) reported no control group (Table 1, Figure 1). Among all studies, 55 (12.5%) included a healthy or uninfected comparator and 14 (3.2%) included another comparator type. One study included both comparator categories. In more granular detail, 29 studies used healthy general population controls, 25 studies used asymptomatic SARS-CoV-2-negative controls (18 by PCR, 5 by serology test, 1 by clinical evaluation and 1 by rapid test), 1 study included patients admitted to hospital settings for conditions entirely unrelated to COVID-19; 9 used retrospective intra-subject designs comparing pre-and post-COVID-19 symptoms, and 5 studies utilized patients presenting with similar respiratory clinical features who ultimately tested negative (4 by PCR or antigen test and 1 by clinical evaluation).

**Table 1.** Methodological characteristics of the evaluated long COVID studies and author-provided supplementary studies.

| Study characteristics | Originally evaluated studies | Studies with additional information from authors | Combined analysis |
| --- | --- | --- | --- |
|  | Number (%) |  |  |
| **Control group status** | **N=440** | **N=19** | **N=459** |
| No control group | 372 (84.5) | 3 (15.8) | 375 (81.7) |
| Healthy/uninfected | 55 (12.5) | 8 (42.1) | 63 (13.7) |
| Other controls | 14 (3.2) | 8 (42.1) | 22 (4.8) |
| **Method of outcome assessment** | **N=440** | **N=19** | **N=459** |
| Solely self-reported | 279 (63.4) | 15 (78.9) | 294 (64.1) |
| Not entirely self-reported | 161 (36.6) | 4 (21.1) | 165 (35.9) |
| **Acknowledgment of uncontrolled design in limitations** | **N=372** | **N=3** | **N=375** |
| No | 252 (67.7) | 3 (100) | 255 (68.0) |
| Yes | 120 (32.3) | 0 (0) | 120 (32.0) |

Note: Comparator categories in control group status were not mutually exclusive. One study included both a healthy/uninfected comparator and another comparator type.

Regarding outcome assessment, 279 (63.4%) studies relied entirely on self-reported symptoms. 161 studies (36.6%) included only clinical evaluation (n=50), laboratory testing (n=14), other non-self-reported assessments (n=20), combined clinical and laboratory testing (n=14), combined clinical and other assessments (n=36), combined laboratory and other assessments (n=1), or utilized all three approaches (n=26).

In 120 of the 372 uncontrolled studies (32.3%), data extractors found some explicit acknowledgement of the absence of a baseline control group in their limitations section. On the remaining 252 studies, we applied a hybrid automated text-mining and manual re-verification approach (Supplementary Material 3). The automated pipelines initially flagged 137 studies for containing control-related terminology, but manual review confirmed that only 8 of them acknowledged the absence of an uninfected baseline comparator outside of their formal limitations paragraph (Supplementary file 4), specifically in the broader Discussion section (n=6) or within the Methods section (n=2). The software initially flagged more studies because the word “control” has other meanings in medical research. Therefore, 244/372 studies (65.6%) did not acknowledge the lack of control limitation anywhere.

### 3.2 Author survey outcomes and data availability

Corresponding authors from 140 (31.8%) of the originally evaluated studies provided a response, leaving 300 (68.2%) classified as non-responders (Table 2). Response rates were similar in authors of studies without control groups (119/372, 32.0%) and in authors of studies who already had control groups in the index systematic review (21/68, 30.9%) (Figure 1).

**Table 2.**
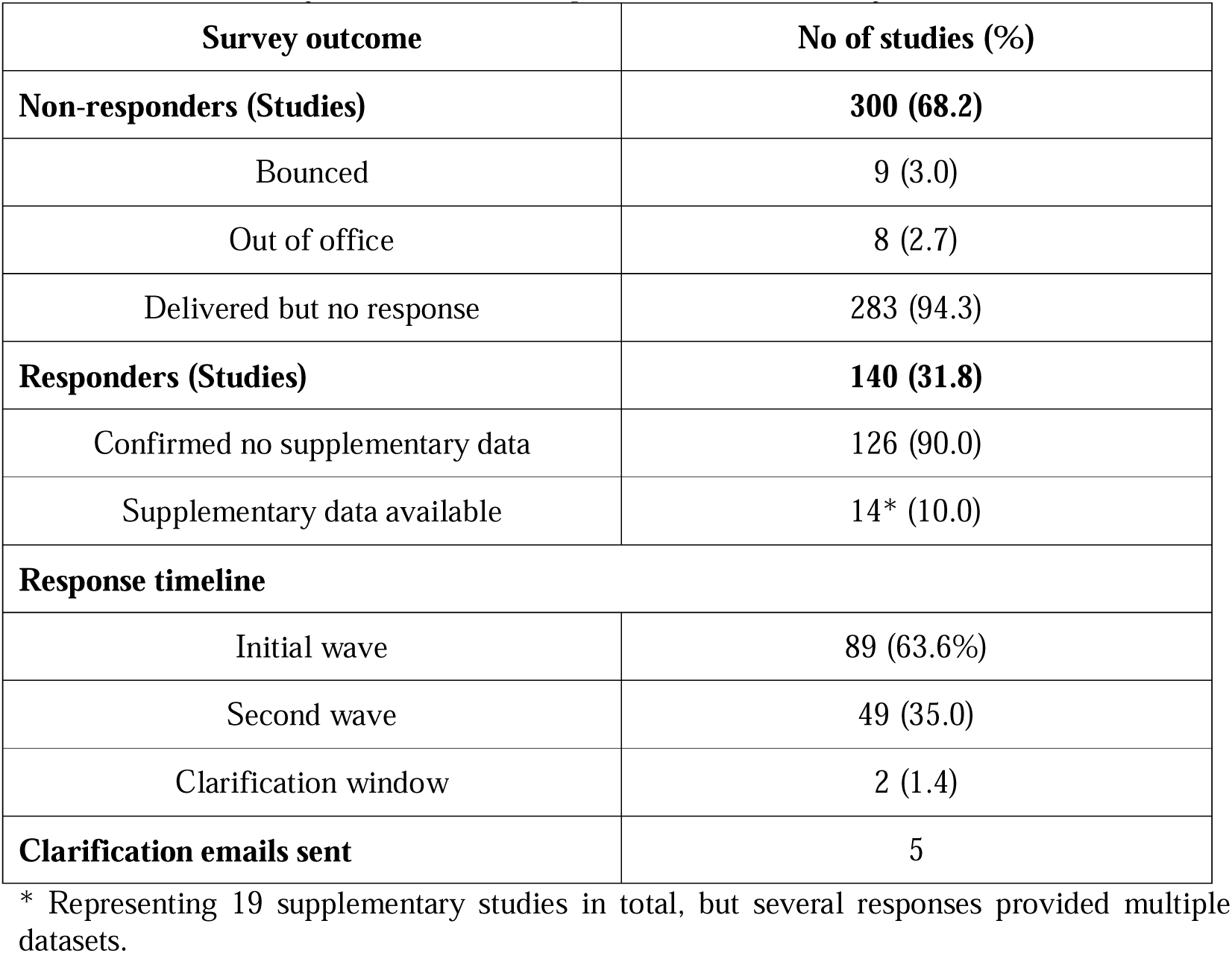
Author survey outcomes and comparative data availability (N = 440).

Among responders, 126 studies (90.0%) did not have any available supplementary comparative data. Supplementary comparative datasets were mentioned for 14 (10.0%) of the original studies. This group includes two respondents who reported having unpublished baseline cohorts (one healthy (under review) and one H1N1 control group) but did not provide further details following clarification requests. 10 directed us to other published studies, one shared a preprint and one mentioned a manuscript currently submitted for peer review. Additional comparative datasets were more likely to be mentioned by authors of studies who already had published other control groups in the index systematic review articles (4/21, 19.0% versus 10/119, 8.4%) (Figure 1).

### 3.3 Additional data and combined analysis

These 14 survey responses ultimately yielded a total of 19 distinct author-provided supplementary datasets (Table 1). Of these 19, 10 were already published by the July 2024 end search date of the index systematic review while the others were published later or were still unpublished. Among these 19, 8 (42.1%) studies incorporated an uninfected control group, 8 (42.1%) utilized other comparators, and 3 (15.8%) lacked controls. Outcome assessment in these datasets used entirely self-reported symptoms in 15/19 (78.9%).

Including these additional datasets, the total number of study reports evaluated was 459 (Table 1) and the overall proportion of studies lacking a control group slightly decreased to 81.7%, while the proportion utilizing healthy and other controls increased to 13.7% and 4.8%, respectively. The prevalence of solely self-reported outcomes remained highly consistent, appearing in 64.1% of the total combined cohort. The overall lack of methodological transparency regarding uncontrolled designs remained largely unchanged, with 68% of uncontrolled studies failing to acknowledge the lack of a baseline control group as a limitation.

In the two months after the closure of pre-specified period for collection of author responses (March 29-May 29, 2026) only 4 additional authors responded, mentioning that no controls were available.

### 3.4 Citation analysis

Controlled studies achieved higher annual citation rates than uncontrolled studies (Median: 7 versus 4, p = 0.002) (Figure 2A). There was no difference between studies using self-reported versus objective assessments (Median: 4 versus 5, p = 0.249) (Figure 2B). Comparing all four design combinations revealed only modest differences (p = 0.012) (Figure 2C) with medians of 8, 7, 4, and 4 for controlled studies with objective assessments, controlled/self-reported, uncontrolled/objective, and uncontrolled/self-reported studies, respectively (Supplementary material 6).

**Figure 2.**
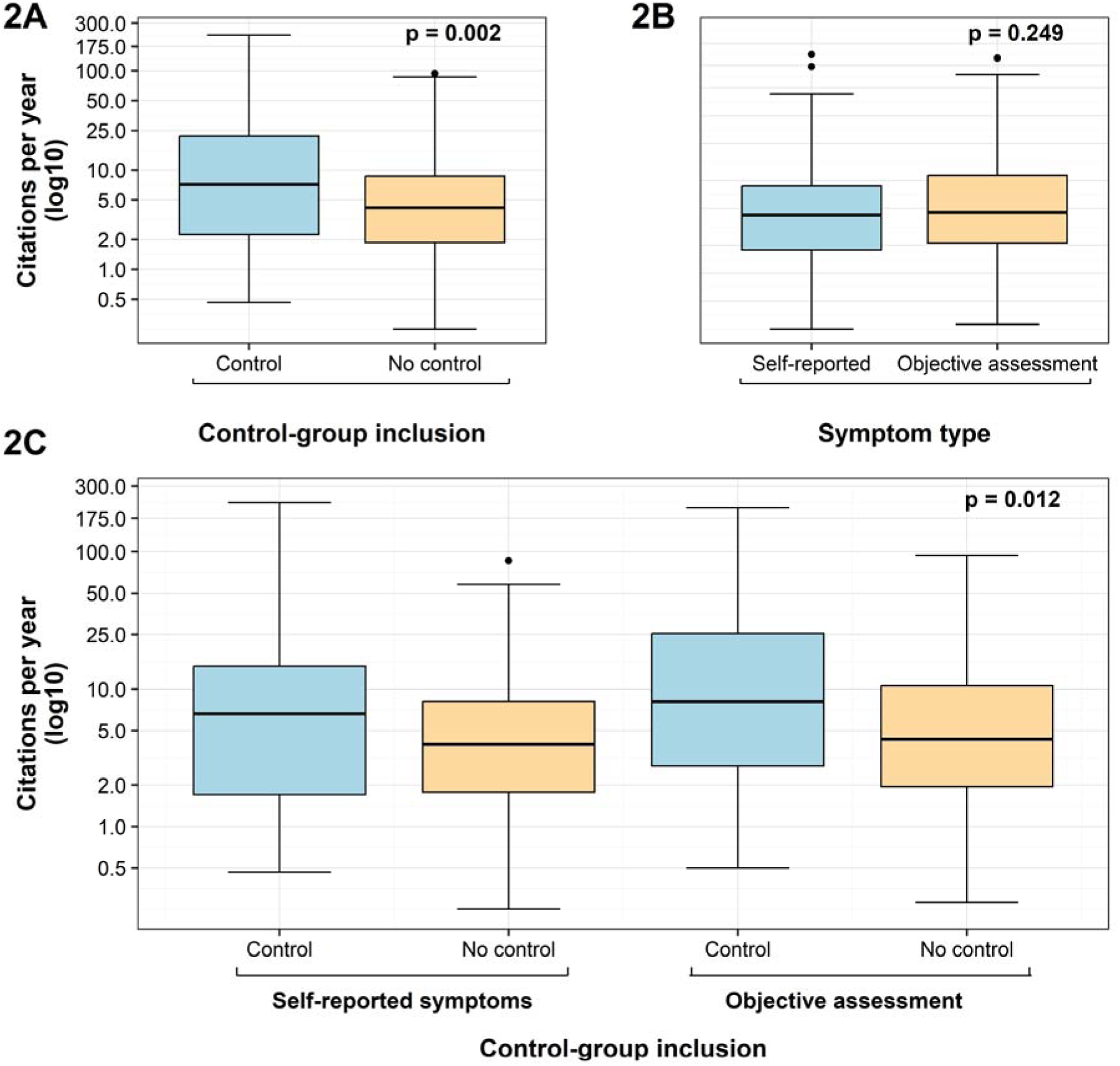
Box plots of number of citations per year for long COVID publications based on A. the inclusion or not of a control group; B. symptoms report; C. both inclusion or not of a control group and type of symptoms report. Boxes represent the interquartile range (IQR), with the horizontal line within the box indicating the median. Whiskers extend to 1.5 times the IQR, and dots represent outliers. Statistical significance was defined as p < 0.005 and p values are shown in the upper right corner of each panel.

## 4. Discussion

The evidence base of long COVID has important fundamental methodological limitations. Our analysis shows that 84.5% of published prevalence studies lacked any baseline comparator, a pattern that persists even after direct author contact, leaving more than four-fifths of the combined analytic cohort without any reported control group. These findings suggest that much of the long COVID prevalence literature is poorly suited to estimate burden attributable specifically to SARS-CoV-2.

This limitation becomes evident when findings from uncontrolled studies are compared with those of controlled studies. Meta-analyses that include studies lacking non-COVID controls have historically generated inflated prevalence estimates suggesting that between 36% and 80% of individuals develop post-acute sequelae [1,9,10]. In pediatric populations, these estimates range between 1% and 25% [11,12]. Reevaluating these clinical outcomes against SARS-CoV-2-negative controls suggests that the relative risk of experiencing at least one persistent symptom is relatively low (RR, 1.37; 95% CI, 1.27-1.49) [13], and even this estimate may be inflated due to residual confounding and selection biases. Uncontrolled observational data may elevate the apparent importance of generic complaints like fatigue and sleep disruption, which controlled studies show are common in the general population rather than unique to the virus [13]. In contrast, indicators of SARS-CoV-2 pathology may be more concentrated in specific impairments such as anosmia, cognitive dysfunction and brain fog [3, 13–15]. Even among these specific impairments, the causal impact of SARS-CoV-2 may vary. E.g. anosmia may have stronger causal documentation, while general neurological and mental manifestations may relate more to or get triggered by other co-existing factors, such as stress and worry [16–19].

Within our combined cohort, 64.1% of investigations relied entirely on self-reported outcomes to establish post-acute sequelae. Such data increase the risk of misclassification. The SARS-CoV-2 pandemic introduced severe societal pressures capable of generating or exacerbating psychological symptoms independently of infection [20]. Quarantine and strict lockdown measures have been associated with elevated anxiety, depressive symptoms, and widespread impairments in memory and daily planning [20–22]. This pandemic-driven psychological distress can skew subjective reporting. Elevated depressive symptoms may lead patients to overestimate their own cognitive dysfunction, resulting in 91% of patients showing a mismatch between reported symptoms and objective reality [23]. Heavy reliance on self-reporting complicates the interpretation of nonspecific symptoms, which may reflect pandemic-and post-pandemic-related distress rather than viral sequelae. Distinguishing infection-attributable symptoms therefore requires comparator data collected in the same time period and setting.

A 31.8% response rate to our author survey aligns with standard expectations [24]. The acquisition of supplementary comparative datasets for only 3.2% of the evaluated studies highlights a reporting gap. Most of these authors supplied data that had already been published elsewhere. While individual studies may have different primary aims, separating prevalence estimates from comparative control data across multiple publications deprives readers of the baseline context needed to interpret morbidity accurately. Implementing best practices for data sharing and methodological transparency is essential to ensure reliability and reproducibility [25].

Two-thirds of the uncontrolled studies evaluated failed to acknowledge the lack of a baseline comparator as a limitation. Lack of reporting of limitations is a persistent problem in the literature beyond just COVID-19 [26] and highlights a structural problem in the peer-review process. During the pandemic, epidemiological standards for comparative inference were often even more difficult to maintain, but the resulting literature should still be interpreted with these design limitations in mind, adding greater caution.

The citation analysis suggested that studies with control groups were modestly more cited, but use of objective symptoms was not associated with higher citation rates. Overall higher citations were not largely driven by better methodological design features. In fact, the most cited original study on long COVID [27] was published before the start date of the search window of the index systematic review [1] and was thus not included in our analysis. That study [27] has received 1906 citations although it had no control group. The most cited article on long COVID [28] with 3718 citations is a non-systematic, expert-based review article that does not discuss at all the importance of proper control groups and simply lists an extremely long list of alleged symptoms and manifestations [28]. The disconnect between methodological rigor and citations has been documented also in other fields [29–31].

Determining which post-acute outcomes are uniquely associated with SARS-CoV-2 necessitates evaluating these sequelae against analogous viral groups. As recently demonstrated, while the choice of diagnostic criteria heavily influences overall prevalence estimates, the rates remain highly comparable between SARS-CoV-2-positive and symptomatic, test-negative cohorts across multiple definitions [33]. Specifically, persistent symptom prevalence at three months ranged from 30.8% to 42.0% following SARS-CoV-2 infection versus 28.1% to 40.3% following other test-negative acute respiratory illnesses, with similar rates also appearing at six months (14.2%–21.9% vs. 14.6%–23.3%) [33].

This diagnostic overlap is further supported by comparative literature suggesting that prolonged symptom clusters and major cardiopulmonary complications may frequently emerge following influenza, RSV, and uncharacterized respiratory infections at equivalent or even higher rates than those observed after COVID-19 [34–36]. Influenza is arguably the most appropriate clinical comparator for SARS-CoV-2 because it is a respiratory virus with a well-documented pandemic history and established population presence and vaccination dynamics.

Our study has some limitations. First, the source cohort relies entirely on the search strategy and inclusion parameters established in the prior index systematic review [1]. Any studies omitted from that database are excluded in our analysis. Second, the initial screening and data extraction for each individual article was performed by a single reviewer. However, we partially mitigated this constraint by automated text mining tools. Third, the targeted email survey is susceptible to non-response bias. Fourth, authors who utilized very unconventional terminology to describe their lack of controls may have still been missed. Fifth, our evaluation focused on the presence and type of comparator groups rather than their full methodological quality. Therefore, studies classified as having comparators may still have been limited by poor matching, residual confounding, or other design weaknesses.

Acknowledging these caveats, research on long COVID should not use uncontrolled prevalence studies to estimate SARS-CoV-2–specific morbidity. Without appropriate comparator groups, such estimates are difficult to interpret and may overestimate the true burden due to the virus. To reduce research waste and establish a more valid evidence base, future studies should apply stricter epidemiological standards, including laboratory confirmation of infection, appropriate matched comparator groups such as patients with influenza or other respiratory infections, and greater use of objective clinical endpoints alongside subjective symptom reports. Proper baseline comparisons are essential for distinguishing post-COVID sequelae from background morbidity and for generating evidence needed to guide patient care.

## Author Contributions (CRediT format)

**Antonios-Periklis Panagiotopoulos:** Conceptualization, Methodology, Investigation, Data curation, Formal analysis, Validation, Project administration, Writing - original draft, Writing - review & editing. **Alexandros Laskaris:** Investigation, Data curation, Validation, Writing - review & editing. **Deny Tsakri:** Investigation, Validation, Writing - review & editing. **Yiannis Manoussopoulos:** Software, Investigation, Data curation, Formal analysis, Writing - review & editing. **Cleo Anastassopoulou:** Validation, Project administration, Writing - review & editing, Supervision. **Athanasios Tsakris:** Writing - review & editing, Supervision. **John P. A. Ioannidis:** Conceptualization, Methodology, Formal analysis, Writing – original draft, Writing - review & editing, Supervision.

## Data access and responsibility

A-P.P. had full access to all the emails received by corresponding authors. All authors had full access to all the other data in the study and take responsibility for the integrity of the data and the accuracy of the data analysis.

## Data Availability

All data produced in the present study are available upon reasonable request to the authors

## Acknowledgments

We thank the corresponding authors who responded to our survey and provided additional information about their studies.

## Supplementary material

Supplementary Table 1. Standardized data extraction fields and study classifications.

Supplementary Material 1. Study protocol and prespecified methodology.

Supplementary Material 2. Email templates

Supplementary Material 3. Informatics pipeline and detailed results.

Supplementary Material 4. Human verification of informatics results.

Supplementary Material 5. Citation information

Supplementary Material 6. Nonparametric citation report

Supplementary Material 7. Complete bibliography of the included original and author-provided studies.

